# Maternal Diet Quality and BMI as Predictors of Human Milk Composition and Exclusive Breastfeeding Duration

**DOI:** 10.1101/2024.09.26.24314257

**Authors:** Hatice Cetinkaya, Christina J. Valentine, Kelly A. Dingess, Nicholas J. Ollberding, Suzanne S. Summer, Nathan A. Meredith, Sarah D. Maria, Ardythe L. Morrow, Laurie A. Nommsen-Rivers

## Abstract

**Background:** Poor diet quality and high body mass index (BMI) contribute to inflammation, which may influence human milk composition and exclusive breastfeeding (EBF) duration.

**Objective:** We evaluated maternal diet and prepregnancy BMI as predictors of human milk C-reactive protein (CRP) and long chain fatty acid concentrations (%LCFA), and EBF duration.

**Methods:** We utilized the Global Exploration of Human Milk Study-Cincinnati subset (n=114), where healthy dyads continued follow-up if ≥75% of feeds were breastmilk at 4 weeks postpartum. We computed a Dietary Inflammatory Index (DII) from diet recalls obtained between 4-13 weeks postpartum, where higher score indicates a more proinflammatory diet. Milk CRP and fatty acid analyses were performed on week 4 milk. We compared milk CRP across 4 combinations of DII×BMI using the Kruskal Wallis test, with BMI categorized as normal versus elevated (<25 versus ≥25 kg/m^2^), and DII split at the median. Linear regression was used to examine DII and BMI as predictors of %LCFA. Logistic regression was used to examine DII tertiles and BMI as predictors of EBF duration.

**Results:** Milk CRP concentrations differed across DII×BMI groups (*p*=0.009): the low DII/normal BMI group had the lowest milk CRP (n=30, median [Q1, Q3], 64.3 [38.2, 121.4] ng/mL) versus all other groups (n=70, 124.1 [71.2, 181] ng/mL, *p*=0.022). Lower milk %LCFA was predicted by higher DII score (β±SE = -0.68 ± 0.21, *p*=0.002, n=103) and higher BMI (β±SE = -0.13 ± 0.01, *p*=0.043, n=114). Having the highest DII tertile and elevated BMI lowered the odds of EBF at week 6 (OR [95% CI]: 0.26 [0.07, 0.85]) compared to the referent group (low or medium DII, normal BMI).

**Conclusions:** Milk CRP concentrations were lowest in women with a more anti-inflammatory diet and normal BMI. Both higher BMI and proinflammatory diet predict lower milk %LCFA and lower EBF prevalence at week 6.

## Introduction

Human milk contains both anti- and pro-inflammatory components, many of which may have biological impact on the breastfeeding infant, such as the regulation of growth, metabolism, immune development, and the microbiome (1, 2). Given that early infancy is a sensitive developmental period (3–5), it is critical to understand the link between modifiable maternal factors and concentrations of milk inflammatory components. Maternal diet and prepregnancy body mass index (BMI) are known to alter some nutrients in milk (6–9), but little is known regarding overall maternal diet quality and BMI in relation to milk concentrations of bioactive compounds.

Both poor diet quality and high BMI contribute to a generalized low-grade inflammatory state (10). Epidemiological studies consistently report an association between obesity and elevated serum concentrations of pro-inflammatory components (11, 12). Moreover, serum concentrations of inflammatory markers are positively associated with consuming a Western-type dietary pattern, as compared to greater adherence to healthy dietary patterns such as the Mediterranean diet (13–16). Elevated inflammatory cytokines in the blood may lead to increased uptake in milk (17), which is consistent with the findings that greater prepregnancy BMI is associated with greater CRP levels in human milk (18, 19). However, our understanding of the effects of maternal diet on human milk inflammatory compounds and its interaction with BMI is limited.

In addition to its effect on milk composition, chronic inflammation may also downregulate milk production (20). Low milk supply is one of the most important reasons for early breastfeeding termination (21, 22). A recent study found that women with very low milk production, despite frequent breastfeeding and/or breast milk expression, had higher levels of milk tumor necrosis factor-alpha (TNF-α) and serum C Reactive Protein (CRP); in addition to displaying evidence of dysfunction in long-chain fatty acids (LCFAs) uptake by the mammary gland. The authors postulated that inflammation suppresses lipoprotein lipase (LPL) activity at the mammary gland, leading to disruption in uptake of LCFAs from blood to milk (20). Given that circulating LCFAs in blood is the predominant source of fatty acids in human milk (23, 24), a diet with pro-inflammatory potential or excessive body weight may lead to dysfunction in milk synthesis as evidenced by lower relative milk LCFAs concentrations (%LCFA), and thereby shorter exclusive breastfeeding (EBF) durations, even in women with high breastfeeding intentions.

The Dietary Inflammation Index (DII) is an evidence-based algorithm to compute the overall inflammation potential of a diet independent of regional or cultural dietary habits (25). A lower DII score indicates a more anti-inflammatory diet, while a higher DII score represents a more proinflammatory diet. DII score is positively associated with higher serum concentrations of inflammatory markers such as CRP, interleukin (IL) - 6, and TNF-α in general adult populations (26–29) and pregnant women (30–32). Current knowledge on the influence of dietary inflammation potential during lactation on milk inflammatory components and breastfeeding outcomes is very limited. Moreover, the existing literature lacks representation of lactating women from the Midwestern U.S., where dietary patterns of general adults tend to be high in energy-dense foods, added sugar, and alcohol, and poor in fiber, vegetables, fruits, whole grains (33), and fish (34). Addressing this knowledge gap is especially timely given that United States Department of Agriculture (USDA) and Department of Health and Human Services recently issued the first detailed dietary guidelines specifically for pregnancy and lactation in the U.S (35). During the extensive review process that led to these guidelines, an advisory committee noted the critical need for more research on the diets of lactating women and its effects on human milk quality to inform future guidelines (36).

Leveraging existing data and biospecimens from a Midwestern cohort of lactating women, we hypothesized that normal prepregnancy BMI and an anti-inflammatory dietary pattern during lactation would be associated with lower human milk CRP levels when compared to participants with an overweight or obese BMI and/or inflammatory dietary pattern. We also examined whether a normal prepregnancy BMI and an anti-inflammatory dietary pattern predicted higher human milk %LCFA and longer EBF duration.

## Methods

### Study Participants

We used de-identified data and biospecimens from the Cincinnati site of the Global Exploration of Human Milk (GEHM) Study (2007–2008) (9, 37), which was a multi-country, longitudinal cohort of mother-infant pairs. Participants were enrolled within the first 2 weeks postpartum and followed up to 52 weeks postpartum. The eligibility of mother-infant dyads was assessed at 2 time points: week 2 and week 4 post birth. Initial selection criteria for women at week 2 were aged between 18 and 49 years, having intention to breastfeed for ≥75% of feeds for ≥3 months, and residing within the Cincinnati metro area. Eligibility criteria for infants at week 2 were being singleton term, and healthy, with a birthweight ≥2500 g. Mothers who did not achieve ≥75% of feeds as breast milk by week 4 were excluded from further follow-up and replaced until a sample size of n=120 eligible dyads was reached. The Institutional Review Board of Cincinnati Children’s Hospital Medical Center approved the GEHM Study, and all mothers provided informed consent in the original study (37). At week 4 of the Cincinnati GEHM study, 120 dyads were deemed eligible to continue with follow-up measurements.

### Primary Exposures: Prepregnancy BMI and Maternal DII score

Maternal sociodemographic and clinical data, including maternal age, race, educational level, and prepregnancy weight and height; and infant birth weight and gestational age, were collected via interview at the baseline GEHM visit. BMI was calculated as weight (kg)/height squared (m^2^). We imputed BMI values for the two participants who had missing height values. To do this, we used the stepAIC() function in R for step-wise variable selection of sociodemographic factors and available anthropometric measurements to predict maternal BMI in the full sample. The final linear regression model included prepregnancy weight, gestational weight gain, and total DII score (R-squared=0.93) in the interpolation of maternal prepregnancy BMI for these two participants.

Maternal dietary data were collected via 24-hour dietary recalls administered on 3 random days between weeks 4 and 13 postpartum during in-person visits or phone interviews. Trained interviewers used the USDA five-step automated multiple-pass method to improve accuracy and portion size instructions were provided to women during the interviews (9). Nutrient analysis was performed using Nutrition Data Systems for Research (NDSR; Nutrition Coordinating Center, University of Minnesota) software, 2013 version. The 3-day average for each dietary component was calculated to obtain estimated usual daily intake.

The DII score for each mother was computed using a combination of the raw nutrient and food group intake data generated in NDSR. The 45 possible dietary parameters of the DII algorithm and our steps in calculating a DII score for each participant are summarized in **Figure 1** and detailed in the **Supplemental Information**; see Shivappa et al. (25) for a detailed explanation of the DII scoring algorithm and how it was derived. The DII score typically ranges from −8.87 (anti-inflammatory) to +7.98 (proinflammatory). There is no strict requirement to use all 45 possible dietary parameters. In our analysis, we included 40 dietary parameters and excluded 5 (alcohol, eugenol, turmeric, saffron, and isoflavones) due to lack of data on these components (see **Figure 1**). Of the 40 included dietary parameters, 33 were individual nutrients and 7 were foods/ingredients (onion, garlic, ginger, pepper, green/black tea, thyme/oregano, and rosemary), all available in NDSR 2013 for assessment of intake (**Supplemental Information**).

**Figure 1.**
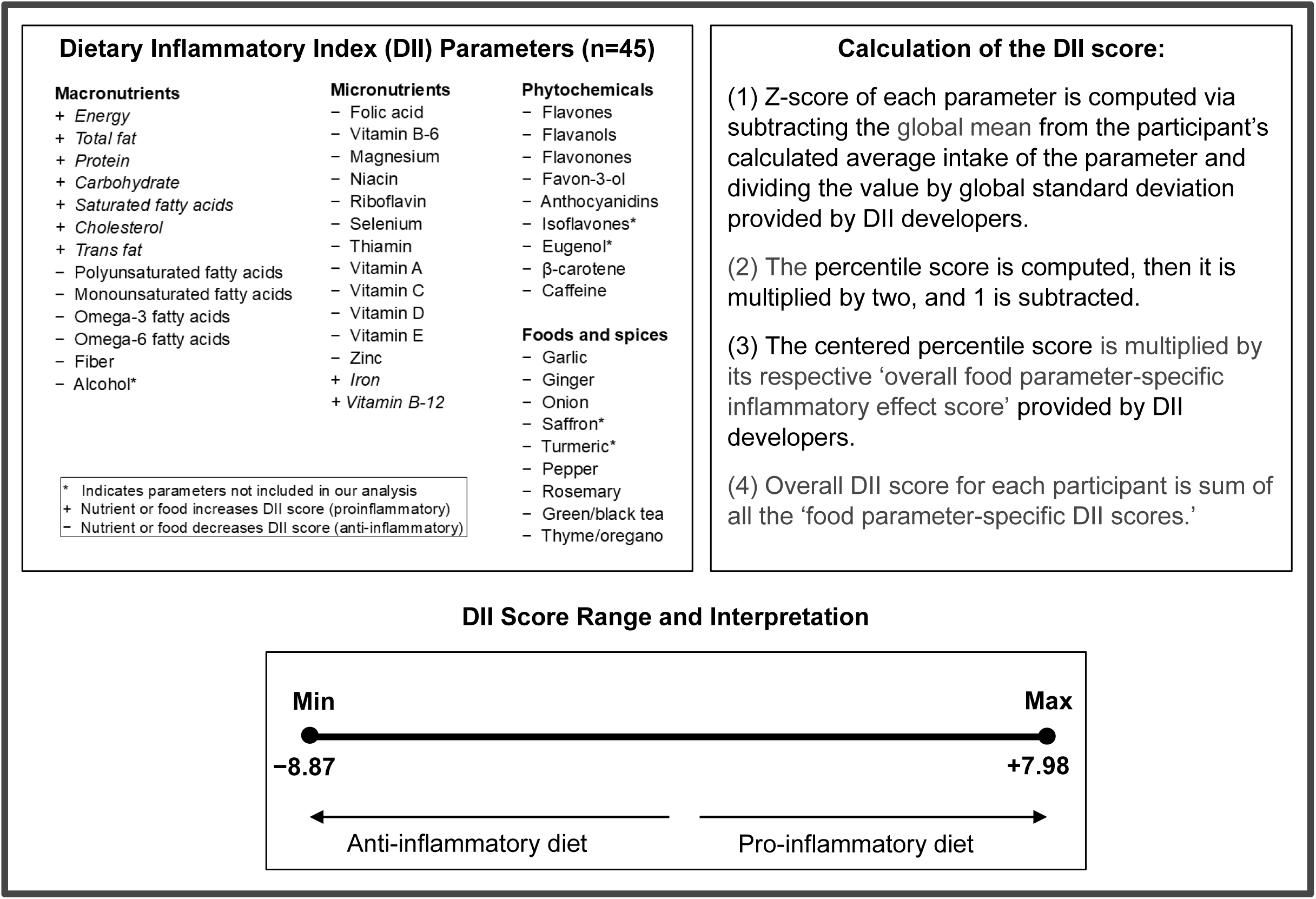
An overview of the Dietary Inflammatory Index (DII) algorithm. DII has 45 parameters and the steps to compute DII score is summarized in this figure. Having a lower DII score indicates having a more anti-inflammatory diet and having a higher DII score indicates having a more pro-inflammatory diet. In our analysis, 40 different parameters were used in the calculation of the DII score.

### Primary Outcome Variable: Quantification of CRP Concentrations in Human Milk

At each in-person visit, mothers provided a milk sample from a complete breast expression using a hospital-grade electric breast pump (Medela Symphony, Medela, Inc., McHenry, Illinois) at a time that was between 9:00am–1:00pm and ≥2 hours since the previous breastfeeding or breast pumping. Collected samples were transferred into labelled 2-mL cryogenic storage vials and stored at −80°C (9).

Prior to quantifying CRP concentrations in human milk, the first author adapted and validated a protocol using a high-sensitivity ELISA kit from R&D Systems (Minneapolis; catalog number DCRP00) to account for human milk rather than serum or urine being the biofluid. Protocols validated for measuring bioactive proteins in serum or urine are often used to quantify these same compounds in human milk, but it is not always known if this practice is valid given the potential matrix effects of human milk on ELISA assay performance (38).

For adaptation and validation of the chosen high-sensitivity CRP ELISA kit, we used the skim fraction of pooled donor milk. First, we generated a standard curve that extended 100% beyond the lower and upper range of the kit to detect the upper and lower limit of quantification of each assay. The standard curve exhibited linearity across the full range of CRP standard dilutions assayed. Then, we determined the optimal dilution factor for human milk. Based on a literature search, the concentration of CRP in human milk is much lower than in serum (39). Thus, we tested assaying milk samples with five different dilution factors (1:99, 1:9, 1:4, 1:3, 1:2, and undiluted), in contrast to the kit’s recommended 100-fold dilution for serum samples. Next, to validate the adapted protocol, we calculated the intra assay precision across 4 replicates and percent recovery of milk samples spiked with a CRP standard across 14 replicates at the final dilution factor. In our ELISA validity experiments, we identified a 1:4 dilution as being the most optimal for assaying CRP in human milk based on CRP concentrations as this dilution most consistently fell along the linear portion of the assay’s standard curve. Dilution factor corrected mean ± SD and coefficient of variation (CV%) for CRP concentration across four replicates of a pooled milk sample assayed at 1:4 dilution was 139.25 ± 2.17 ng/mL and 1.56%, respectively. The spike recovery of CRP in human milk ranged from 89.9% to 120.2% across 14 replicates, with a CV% of 4.27% (**Supplemental Tables 1 and 2**).

Once we validated our adapted protocol for assay of milk CRP, we used the same kit to assay week 4 milk samples in duplicate. Whole milk samples collected at week 4 were thawed, centrifuged at 4,000xg for 10 minutes at 4°C, and the milk fat layer removed to use skim milk for the CRP assay.

### Secondary Outcome Variables: Milk Fatty Acid Concentrations and EBF Duration

Methodology for the milk fatty acid analysis of the GEHM study samples has been reported previously (9). Briefly, total lipids were extracted from whole milk based on the modified Bligh and Dyer method (40), using an Agilent 7890 gas chromatograph. For the purposes of the present analysis, fatty acids were categorized into 3 groups based on carbon chain length: medium chain fatty acid (MCFAs, <16 carbons), 16-carbon fatty acids, and LCFAs (>16 carbons), and presented as the relative percentage (weight%) of total fatty acid concentration. This categorization enables examination of the source of fatty acids in the formation of milk lipids, as LCFAs are derived from the bloodstream, while MCFAs are synthesized by the mammary gland with de novo lipogenesis, and 16-carbon fatty acids are derived from both exogenous and endogenous sources (23, 24).

In the Cincinnati GEHM study, breastfeeding status was determined through weekly telephone surveillance (37). According to the World Health Organization, duration of EBF is defined as the age of the infant at the last time only breast milk was provided prior to the introduction of any complementary foods or drinks, including water (41). Based on whether the dyad met this definition, we coded EBF status (yes/no) at 6 and 13 weeks postpartum.

### Statistical Analysis

Participant demographic and clinical variables were presented using means ± standard deviations (SD) or medians [Quartile 1, Quartile 3] ([Q1, Q3]) for continuous variables and frequencies (proportion) for categorical variables. One way ANOVA or Kruskal Wallis tests and Chi-square or Fisher exact tests (when cell sizes ≤5) were used to test for differences across DII and BMI groups.

To test our primary hypothesis, we compared milk CRP levels across four combinations of DII and BMI groups. For this analysis, participants were categorized into low and high DII groups at the median DII score for the sample, and categorized as normal (<25 kg/m^2^) or elevated (≥25 kg/m^2^) BMI based on the WHO cut-off for healthy BMI (42). CRP levels were compared using the Kruskal Wallis test, followed by post-hoc analyses of comparing the group with low DII/normal BMI versus the others with the Kruskal Wallis test, and pair-wise comparisons using Dunn’s test with Bonferroni correction.

Linear regression was used to determine whether maternal DII or prepregnancy BMI were associated with milk fatty acid profiles. Bivariate associations of maternal DII score and prepregnancy BMI were correlated with the relative concentrations of each fatty acid group with all variables modeled as continuous terms. We used all available data for each analysis to maximize power.

Given that individual LCFAs consumed in the diet, particularly omega-3 and omega-6 fatty acids, are highly correlated with their relative concentrations in human milk (43–45), we performed a sensitivity analysis to determine the extent to which the above linear regression results were influenced by fatty acid parameters in the DII scoring algorithm. To do this, we re-computed the total DII score after removing PUFA, omega-3 fatty acids, and omega-6 fatty acids from the scoring algorithm and repeated the above linear regression analysis.

To examine the joint association of dietary pattern and BMI with EBF duration, we first compared EBF status at weeks 6 and 13 postpartum across maternal and infant characteristics to identify potential confounding variables. Next, the duration of EBF across DII tertiles was illustrated with Kaplan Meier survival curves up to week 13 postpartum. Since survival analysis revealed nearly identical curves for the lower two DII tertiles, these tertiles were combined to maximize power in further analyses (low or medium DII versus high DII**, Supplemental Figure 1**). Then, we conducted logistic regression analysis to determine if DII or BMI category (normal versus above normal) predicted EBF status at week 6 or 13, with adjustment for characteristics where the bivariate association with EBF exhibited a p-value <0.20. Results for the crude and adjusted regression model are reported as odds ratio [95% confidence interval] (OR [95% CI]) and adjusted OR [95% CI] (aOR [95% CI]), respectively.

Data analysis was performed using the R version 4.2.2 (46). The MASS package (version 7.3.60.0.1) was used to perform variable selection for missing data imputation (47). The glmnet package (version 4.1.8) was used to conduct logistic regression models (48).

## Results

### Participant Characteristics

Within the Cincinnati subcohort of the GEHM Study a total of 120 mother-infant dyads were eligible for continued follow-up at week 4 postpartum, including meeting the criterion of providing ≥75% of feeds as breast milk. A summary of the available sample sizes for each analysis is provided in **Figure 2**. The DII score ranged from -3.37 to 7.02, with a median [Q1, Q3] of 0.19 [-1.50, 1.27] units. Mean (± SD) maternal age was 31.7 ± 5.1 years at delivery, and median [Q1, Q3] prepregnancy BMI was 25.6 [22.8, 31.1] kg/m^2^. The proportion of Black and White women in our sample were 12.0% and 86.1%, respectively, with 1.9% Native Hawaiian or other Pacific Islander (**Supplemental Table 3**). **Table 1** summarizes maternal and infant characteristics stratified by DII and BMI groups. Prepregnancy BMI and total DII score differed significantly across the four combinations of DII and BMI groupings, but no other maternal or infant characteristic reached statistical significance when compared across the four groups.

**Figure 2.**
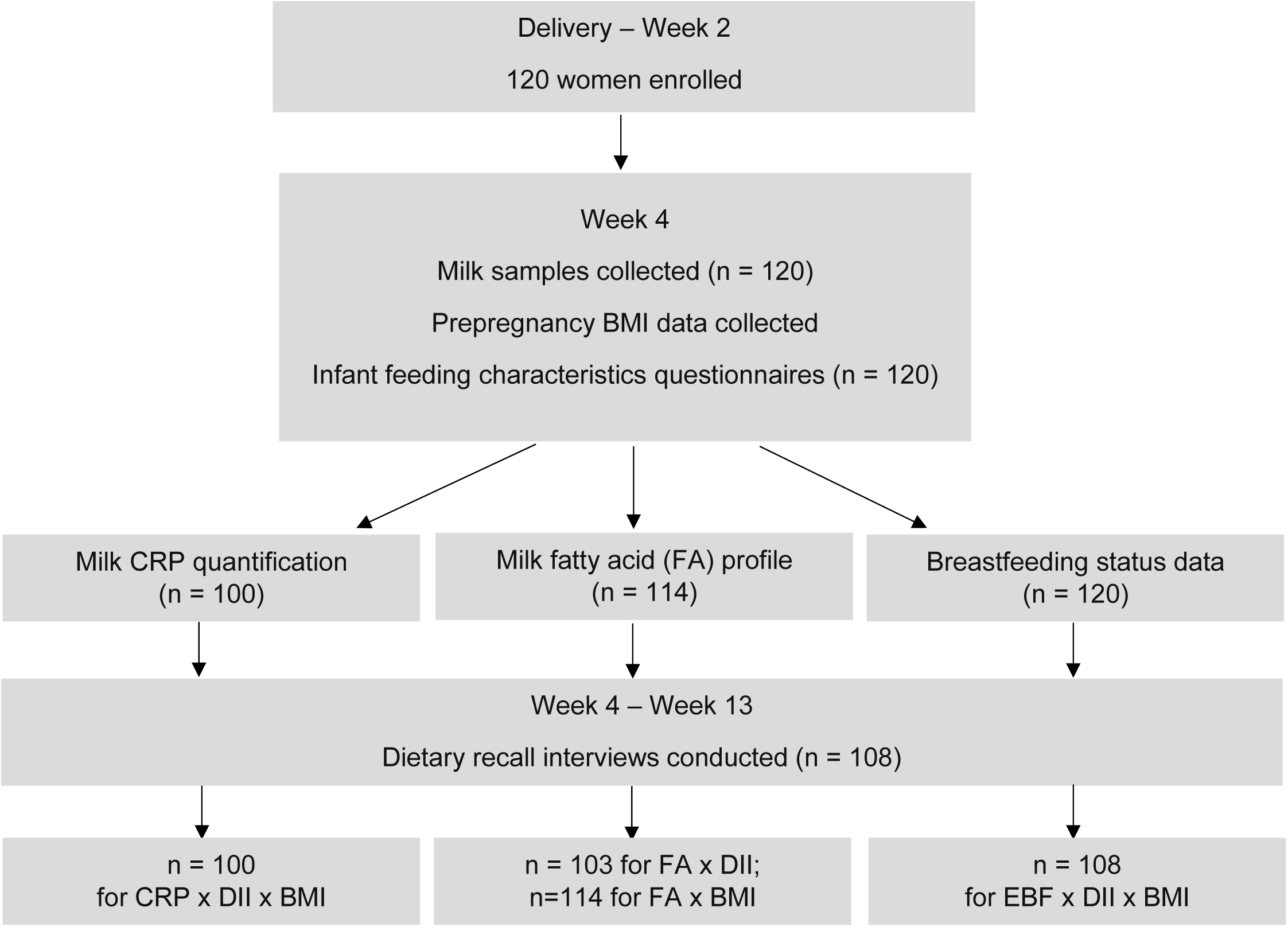
Flowchart for the Cincinnati site of the Global Exploration of Human Milk (GEHM) Study participants. Of 120 mother-infant dyads deemed eligible for follow-up at week 4 postpartum, maternal dietary data was collected for n = 108, breastfeeding status and prepregnancy BMI were collected for n = 120, and C-reactive protein (CRP) quantification and fatty acid profile of human milk samples collected at week 4 were performed for n = 100 and n = 114, respectively.

**Table 1.** Summary of maternal and infant characteristics across DII and BMI groups^1^.

| Maternal and Infant Characteristics<sup>2</sup> | Low DII/Normal BMI<br>(n = 32) | Low DII/Elevated BMI<br>(n = 22) | High DII/Normal BMI<br>(n = 17) | High DII/Elevated BMI<br>(n = 37) | p value |
| --- | --- | --- | --- | --- | --- |
| Total DII score | -1.6 [-2.2, -0.8] | -1.4 [-2.3, -0.3] | 1.5 [1.1, 1.9] | 1.2 [0.5, 2.3] | < 0.001 |
| Prepregnancy BMI, kg/m<sup>2</sup> | 22.2 [21.2, 23.3] | 30.5 [27.0, 35.6] | 23.3 [21.5, 24.2] | 30.1 [27.3, 33.1] | < 0.001 |
| Maternal delivery age, y | 32.0 ± 5.1 | 30.7 ± 4.2 | 30.9 ± 4.5 | 32.5 ± 5.8 | 0.52 |
| Maternal race |  |  |  |  | 0.06 |
| Black | 3 (9.4) | 0 (0) | 2 (11.8) | 8 (21.6) |  |
| White | 28 (87.5) | 22 (100) | 14 (82.3) | 29 (78.4) |  |
| Others | 1 (3.1) | 0 (0) | 1 (5.9) | 0 |  |
| Maternal education |  |  |  |  | 0.19 |
| High school graduate | 0 (0) | 2 (9.1) | 1 (5.9) | 2 (5.4) |  |
| College degree | 26 (81.2) | 15 (68.2) | 11 (64.7) | 32 (86.5) |  |
| Post-college education | 6 (18.8) | 5 (22.7) | 5 (29.4) | 3 (8.1) |  |
| EBF duration, d | 113.5 [70.0, 150.0] | 91.5 [61.3, 120.5] | 100.0 [47.0, 163.0] | 92.0 [32.0, 123.0] | 0.27 |
| BF duration, d | 377.0 [236.3, 541.8] | 338.0 [180.5, 380.8] | 364.0 [170.0, 403.0] | 330.0 [151.0, 689.0] | 0.16 |
| Infant sex, male | 14 (43.8) | 12 (54.5) | 10 (58.8) | 14 (37.8) | 0.42 |
| Infant gestational age, wk | 39.7 ± 1.1 | 39.6 ± 0.9 | 39.3 ± 1.1 | 39.3 ± 0.9 | 0.25 |
| Infant birthweight, kg | 3.6 ± 0.5 | 3.7 ± 0.4 | 3.5 ± 0.4 | 3.6 ± 0.4 | 0.74 |

<sup>1</sup> Participants (n = 108) were categorized into low and high dietary inflammatory index (DII) groups at the median and body mass index (BMI) groups defined as normal (<25 kg/m<sup>2</sup>) or elevated (≥25 kg/m<sup>2</sup>).
<sup>2</sup> Values are mean ± SD or median [Quartile 1, Quartile 3] for parametric and non-parametric continuous variables, respectively or n (%) for frequency (proportion) of categorical variables, n = 108. Based on the normality of data, differences in continuous variables
across the groups were compared with either one-way ANOVA test or Kruskal Wallis test. Proportions of categorical variables across the DII and BMI groups were compared with Pearson chi-square test or Fisher exact tests in instances with cell sizes $\leq 5$ . kg/m<sup>2</sup>, kilograms per meter square; y, years; d, days; wk, weeks

### Human Milk CRP Concentrations

For the primary aim, analysis included n = 100 mothers with both milk CRP and DII results. Within this subsample, human milk CRP levels ranged from 3.1 to 477.4 ng/mL, with a median [Q1, Q3] of 101.7 [54.8, 172.3] ng/mL. Human milk CRP concentrations were significantly different across the four categories of DII and BMI groups (*p* = 0.009, **Table 2**). In post-hoc analysis, women with low DII/normal BMI had significantly lower milk CRP (n = 30, 64.3 [38.2, 121.4] ng/mL) as compared to the other three groups combined (n = 70, 124.1 [71.2, 181] ng/mL, *p* = 0.022). CRP levels for those with low DII/elevated BMI (n = 20), high DII/normal BMI (n = 17), and high DII/elevated BMI (n = 32) were 152.4 [96.4, 205.9], 110.4 [82.5, 163.2], and 107.7 [54.3, 186.1] ng/mL, respectively (**Table 2**). Pairwise comparisons revealed that women with both low DII and normal BMI had significantly lower milk CRP concentrations than each of the other three groups, but the only significant pair-wise comparison after Bonferroni adjustment was low DII/normal BMI group versus low DII/elevated BMI group (adjusted *p=* 0.007, **Figure 3**). Multivariable linear regression analysis showed that each one unit increase in prepregnancy BMI was associated with 5.25 ng/mL increase in CRP levels in milk, while DII was not a significant predictor of milk CRP concentrations (**Supplemental Table 4**).

**Figure 3.**
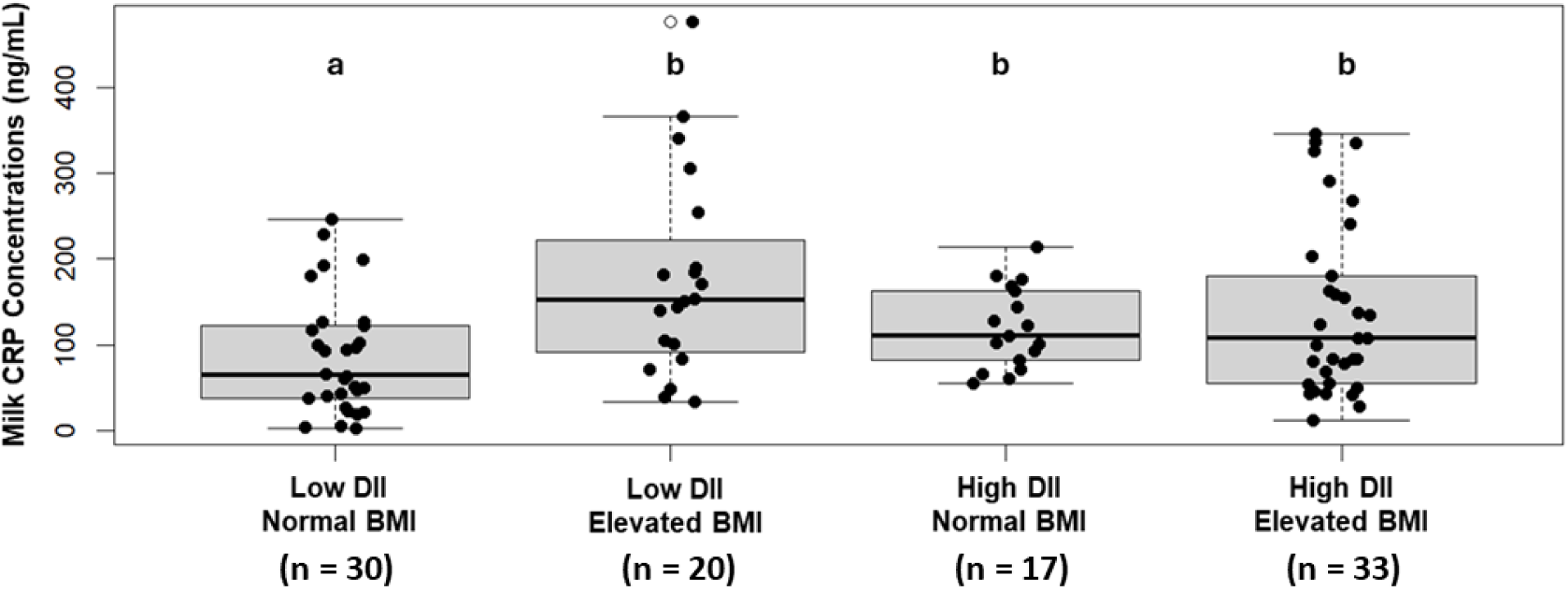
Box plots showing medians and quartiles of milk CRP concentrations (ng/mL) by DII and BMI groups. Differences in milk CRP levels were compared usng the Dunn’s test with Bonferroni adjustment. Differing letters above each group denotes significant difference prior to Bonferroni adjustment (p<0.05). The group with both low DII and normal BMI had the lowest milk CRP levels, but after Bonferroni adjustment it was only significantly different from the group with low DII and elevated BMI (adjusted *p*= 0.007). DII, Dietary Inflammatory Index; BMI, body mass index.

**Table 2.** Comparison of c-reactive protein (CRP) concentrations in human milk across DII and BMI groups^1^.

| Comparison across all four DII × BMI groups<sup>2</sup> |  |  |  | Post-hoc analysis<sup>2,3</sup> |  |  |
| --- | --- | --- | --- | --- | --- | --- |
| DII × BMI category | n (%) | Milk CRP (ng/mL)<br>Median [Q1, Q3] | p-value | n (%) | Milk CRP (ng/mL)<br>Median [Q1, Q3] | p-value |
| Low DII<br>Normal BMI | 30 (30) | 64.3 [38.2, 121.4] | 0.0096 | 30 (30) | 64.3 [38.2, 121.4] | 0.0022 |
| Low DII<br>Elevated BMI | 20 (20) | 152.4 [96.4, 205.9] |  | 70 (70) | 123.1 [73.0, 180.9] |  |
| High DII<br>Elevated BMI | 17 (17) | 110.4 [82.5, 163.2] |  |  |  |  |
| High DII<br>Elevated BMI | 33 (33) | 107.5 [54.7, 180.4] |  |  |  |  |

<sup>1</sup> Participants (n = 100) were categorized into low and high dietary inflammation index (DII) groups at the median and body mass index (BMI) groups defined as normal (<25 kg/m<sup>2</sup>) or elevated (≥25 kg/m<sup>2</sup>); n (%), frequency (proportion); Median [Q1, Q3], median [Quartile 1, Quartile 3]
<sup>2</sup> Group differences were compared with the Kruskal Wallis test.
<sup>3</sup> For post-hoc analysis, milk CRP levels in women with both low DII score and normal BMI (n = 30) were compared with the other three categories combined (n = 70).

### Human Milk Fatty Acid Composition

Among the n = 114 with human milk fatty acid data, mean ± SD relative concentrations of MCFA, 16-carbon fatty acids, and LCFA were 12.1 ± 3.7, 22.5 ± 2.9, and 65.4 ± 4.6%, respectively. A higher DII score was significantly associated with lower %LCFA (β ± SE = -0.68 ± 0.21, *p* = 0.002, n = 103) and higher %MCFA (β ± SE = 0.49 ± 0.16, *p* = 0.003, n = 103) in human milk. Also, higher prepregnancy BMI significantly predicted lower milk %LCFA (β ± SE = -0.13 ± 0.06, *p* = 0.043, n = 114). No association was found between relative concentrations of 16-carbon fatty acids in milk and DII score or prepregnancy BMI (**Table 4**). In sensitivity analysis, the association between maternal DII score and milk %LCFA remained significant when using a DII score that was calculated after the exclusion of LCFA related dietary parameters (**Supplemental Table 5**).

### EBF Outcomes at Week 6 and 13 Postpartum

As mentioned previously, to be eligible for continued follow-up in the original GEHM study, >=75% of feeds had to be breast milk at 4 weeks postpartum. Among the dyads deemed eligible for continued follow-up at week 4 postpartum, 78.7% and 59.3% achieved EBF to 6 and 13 weeks, respectively. **Table 3** summarizes maternal and infant characteristics comparing EBF status at weeks 6 and 13. At both weeks 6 and 13, total DII score and prepregnancy BMI were significantly different according to EBF status at these time points. To determine the joint associations of DII and BMI with EBF status, we modeled EBF status stratified by DII group (low or medium versus high) and prepregnancy BMI (normal versus elevated) with adjustment for maternal race and age. As illustrated in **Figure 4**, the high DII/elevated BMI group had a significantly lower prevalence of achieving EBF to week 6 as compared to the referent group (low or medium DII/normal BMI, aOR [95% CI]: 0.28 [0.07, 097]), but there was no significant difference between any of the three DII and BMI groups as compared to the referent group at week 13.

**Figure 4.**
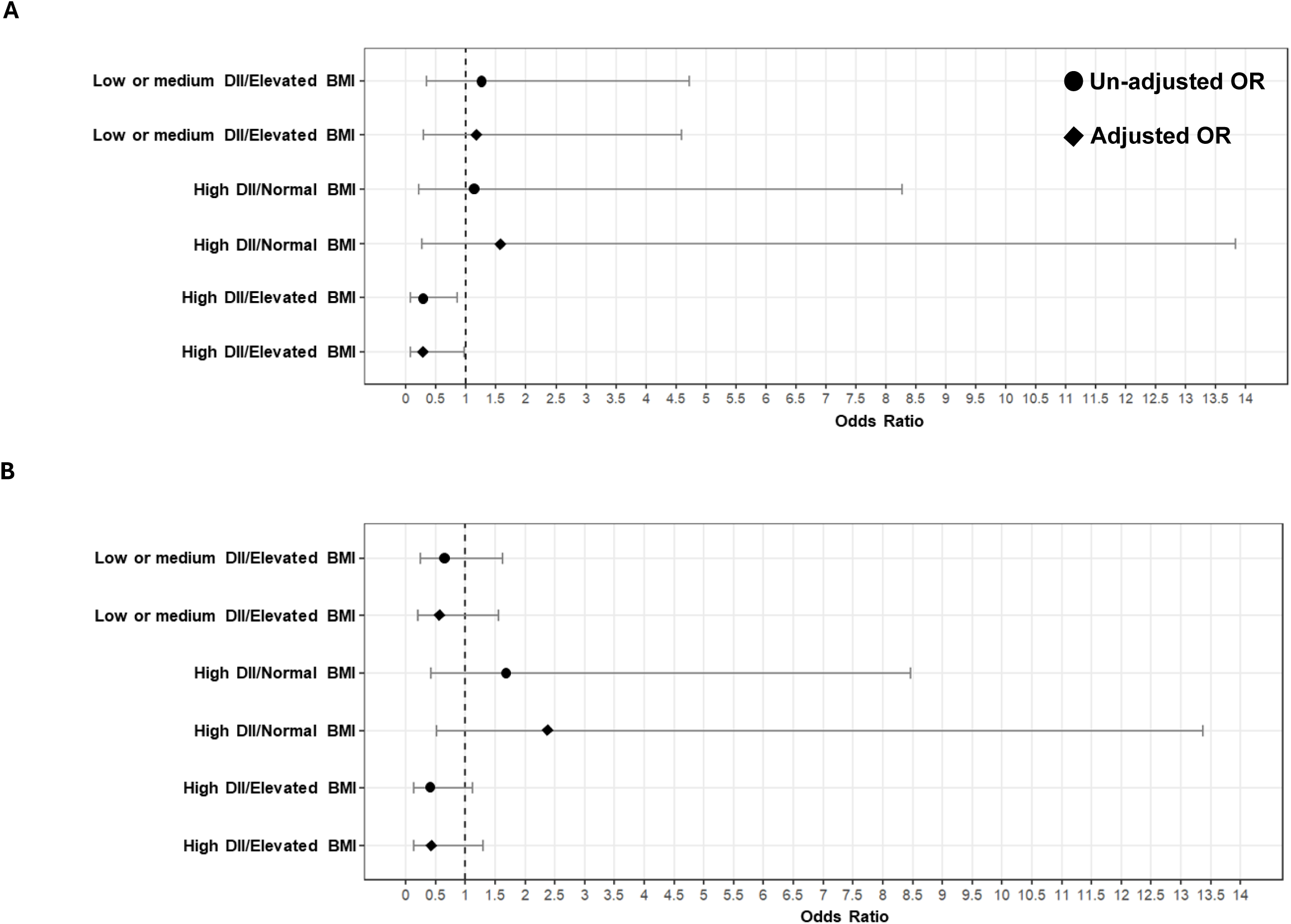
Joint associations of dietary inflammation index (DII) and body mass index (BMI) with the odds of exclusive breastfeeding (EBF) to weeks (A) and week 13 (B) postpartum. Only women with high DII and elevated BMI had lower odds of continuing EBF at week 6 (OR [95% CI]: 0.26 [0.07, 0.85]) compared to the referent group (low/medium DII, normal BMI); this association remained significant after adjusting for maternal race and age (aOR [95% CI]: 0.28 [0.07, 0.97]) Both crude and adjusted odds ratios [95% confidence interval] (OR [95% CI]) are illustrated by DII and BMI group: low/medium DII / elevated BMI (n = 36), high DII / normal BMI (n = 13), and high DII / elevated BMI (n = 23) in comparison to women with low/medium DII / normal BMI (reference group, n = 36). The model was adjusted for maternal race and maternal age.

**Table 3.** Maternal and infant characteristics by exclusive breastfeeding (EBF) outcomes at weeks 6 and 13 postpartum.

| Maternal and infant characteristics<sup>1</sup> | At 6 weeks |  |  | At 13 weeks |  |  |
| --- | --- | --- | --- | --- | --- | --- |
|  | EBF (n = 85) | No EBF (n = 23) | p-value | EBF (n = 64) | No EBF (n = 44) | p-value |
| Total DII score | 0.1 [-1.7, 1.1] | 1.2 [-0.6, 2.4] | 0.0005 | 0.2 [-1.8, 1.1] | 0.3 [-1.1, 1.4] | 0.03 |
| Prepregnancy BMI, kg/m<sup>2</sup> | 25.2 [22.4, 30.9] | 27.2 [24.4, 31.7] | <0.0001 | 24.4 [22.0, 30.5] | 26.7 [24.2, 31.6] | <0.0001 |
| Maternal race |  |  |  |  |  |  |
| Black | 6 (7.1) | 7 (30.4) | 0.003 | 3 (4.7) | 10 (22.7) | 0.01 |
| White | 78 (91.8) | 15 (65.2) |  | 60 (93.7) | 33 (75.0) |  |
| Others | 1 (1.1) | 1 (4.4) |  | 1 (1.6) | 1 (2.3) |  |
| Infant sex, male | 39 (45.9) | 11 (47.8) | 1 | 35 (54.7) | 15 (34.1) | 0.056 |
| Maternal education |  |  |  |  |  |  |
| High school graduate | 4 (4.7) | 1 (4.4) | 1 | 3 (4.7) | 2 (4.6) | 0.90 |
| College degree | 15 (17.6) | 18 (78.3) |  | 49 (76.5) | 35 (79.5) |  |
| Post-college education | 66 (77.7) | 4 (17.3) |  | 12 (18.8) | 7 (15.9) |  |
| Maternal age, years | 31.9 ± 4.9 | 31.2 ± 6.1 | 0.17 | 32.3 ± 5.1 | 30.90 ± 5.0 | 0.13 |
| WIC enrollment post-birth, no. (%) yes | 18 (21.2) | 8 (34.8) | 0.281 | 13 (20.3) | 13 (29.5) | 0.38 |

<sup>1</sup> Values are mean ± SD or median [Quartile 1, Quartile 3] for parametric and non-parametric continuous variables, respectively, or n (%) for frequency (proportion) of categorical variables, n = 108. EBF, exclusive breastfeeding; DII, Dietary Inflammatory Index; BMI, body mass index; WIC, Women, Infants, and Children Supplemental Food Program. Independent t-test or Mann-Whitney U-test was used for continuous variables with parametric and non-parametric distribution, respectively. Pearson chi-square test or Fisher exact tests (in instances with cell sizes ≤5) was used to compare the proportions of categorical variables across EBF outcome groups.

**Table 4.** Associations of maternal DII score and prepregnancy BMI with relative concentrations of milk fatty acids at week 4 postpartum^1^.

| Milk fatty acid group<sup>2</sup> | Relative % <sup>3</sup><br>(Mean ± SD) | Association with DII score<br>(n = 103) |  | Association with prepregnancy BMI<br>(n = 114) |  |
| --- | --- | --- | --- | --- | --- |
| | | $\beta \pm SE$ | *p* value | $\beta \pm SE$ | *p* value |
| MCFA | 12.1 ± 3.7 | 0.49 ± 0.16 | 0.003 | 0.09 ± 0.05 | 0.074 |
| C16 | 22.5 ± 2.9 | 0.19 ± 0.14 | 0.192 | 0.04 ± 0.04 | 0.363 |
| LCFA | 65.4 ± 4.6 | -0.68 ± 0.21 | 0.002 | -0.13 ± 0.06 | 0.043 |

<sup>1</sup> Associations of maternal dietary inflammatory index (DII) score and preprenancy body mass index (BMI) with relative fatty acid concentrations in human milk were modeled using linear regression. To maximize power, we used the maximum available data for each analysis. Data presented are beta coefficient ± standard error ( $\beta \pm SE$ ).
<sup>2</sup> Fatty acid groups were created based on fatty acid carbon chain length: medium chain fatty acids (MCFA) are < 16 carbons, C16 = 16 carbon fatty acids, and long chain fatty acids (LCFA) are > 16 carbons.
<sup>3</sup> Relative concentrations of each milk fatty acid group computed based on their percent of total milk fatty acids.

## Discussion

This study examined the joint association of maternal dietary inflammation potential and prepregnancy BMI with milk CRP, milk fatty acid levels, and EBF status in a Midwestern cohort. We employed DII to compute the inflammatory potential of maternal diet. DII score in our study ranged from -3.37 to +7.02. Based on more than 160 papers published between 2014 and 2018, Herbert et al. (49) reported that the DII score generally ranges from −5.5 to +5.5 when derived from 25-30 parameters. Our analysis used 40 parameters, and our findings align with the effective range of DII provided by its developers (49), which indicates that our method for scoring is valid. The CRP concentrations in the week 4 GEHM milk samples reported here - median of 101.7 ng/mL with an interquartile range of 54.8 to 172.3 – indicates a higher concentration and much wider range of human milk CRP than previously reported (18, 19). For example, Whitaker et al. reports a median CRP of 24.5 and an interquartile range of 11.2 to 67.1 ng/mL for week 4 samples using an ELISA assay (19). While differences may be explained by analysis of different populations, we speculate that our wider range may be explained by being able to quantify milk CRP at the edges of the limit of detection by using a high-sensitivity ELISA kit that was validated and optimized for human milk.

In our study, we found that mothers with an anti-inflammatory dietary pattern and prepregnancy BMI < 25 kg/m^2^ had the lowest milk CRP levels compared to the other three combinations of DII and BMI. Our findings align with two previous studies that reported a positive association between maternal prepregnancy BMI and milk CRP concentrations (18, 19). However, to the best of our knowledge, the role of maternal diet quality on milk CRP concentrations has not been previously published. Ramiro-Cortijo et al. (50) found no significant association between postpartum DII score and other cytokines in human milk, including IL-8, IL-10, IL-13, and TNF-α, but did not study CRP. However, the Ramiro-Cortijo study (50) employed only 20 parameters for DII scoring, which may have limited their ability to capture dietary inflammatory potential, and they did not report whether their assay protocols were validated for use in human milk. Aside from having only 35 dyads with milk cytokine assessment, the potential interaction of maternal BMI with diet quality was not considered. In the present study, we observed the modifying effects of diet on the association of prepregnancy BMI with CRP levels in human milk.

Elevated CRP levels in the milk of mothers with elevated BMI and/or a pro-inflammatory dietary pattern can be explained by higher uptake of CRP from circulation by the mammary glands among mothers with higher blood CRP levels. Because obesity can lead to chronic systemic inflammation due to increased release of pro-inflammatory cytokines such as TNF-α and IL-6 from adipose tissue, which induces synthesis of CRP in hepatocytes (11). Furthermore, a poor-quality diet may upregulate inflammatory responses via increasing oxidative stress, dysbiosis of gut microbiota, and worsening glycemic control (51). Given that poor diet quality or elevated BMI can lead to increases in inflammatory markers in circulation (12, 14, 15), we may speculate that mothers with high diet quality and BMI < 25 kg/m^2^ might have lower serum CRP levels than others. However, we could not assess serum CRP levels, and the correlation of CRP levels between serum and human milk remains unclear. While the correlation between DII score and serum concentrations of cytokines has been demonstrated in prior studies of non-lactating populations (26–29), little is known about the effects of maternal dietary inflammation potential on milk inflammatory components. Given the critical need for more research on the association between maternal diet and human milk composition to inform future guidelines, we believe our study contributes to the existing literature by being the first cohort study to investigate the interplay between maternal diet quality and BMI on milk CRP levels.

The biological impacts of human milk inflammatory markers on breastfed infants are not yet known. Given that human milk contains both proteases and antiproteases, one can infer that proteases may assist the digestion of proteins in the breastfeeding infant’s immature digestive system, while antiproteases may have evolved to protect the bioactive compounds in human milk from digestion (52). Although the mechanistic studies that investigate the bioavailability of these bioactive compounds in infant’s digestive system are lacking, a growing number of studies have shown the potential effects of some milk inflammatory markers on infant growth outcomes (53). Particularly, Sims et al. (18) showed that greater intake of CRP via breastfeeding during the first 9 months of life is positively associated with greater fat mass index in infants of mothers with prepregnancy BMI ≥25 kg/m^2^, while it was negatively associated with fat free mass in the infants of mothers with normal prepregnancy BMI. However, our understanding of the long-term effects of CRP intake on the breastfed infants is limited.

In study mothers, 78.7% and 59.3% continued EBF to 6 and 13 weeks, respectively, which is higher than regional and national statistics (54). This difference can be explained by the enrollment criteria of the GEHM Study, which selected for mothers with high breastfeeding intention and success (37). Comparing DII and BMI groups, we found a significant association with EBF at week 6, and a trend but not a significant difference in EBF at week 13, and speculate that sociodemographic and social factors, including return to work, are increasingly important contributors to cessation of EBF at later stages of lactation.

Both higher dietary inflammation potential and prepregnancy BMI were associated with lower %LCFA in milk and lower week 6 EBF duration in the present study. A recent analysis of three international cohorts (n = 5120 dyads) demonstrated that the inverse association between maternal obesity and duration of EBF and significantly mediated by higher maternal DII score during pregnancy (55). Our findings support the hypothesis that systemic inflammation may hamper breastfeeding (20). While we acknowledge that breastfeeding duration has multiple determinants that were not addressed in this study (56), physiological factors may contribute to shortened EBF duration in the early postpartum period and may be particularly apparent in women with strong breastfeeding intentions, such as those included in our study. For example, one study reported a higher prevalence of elevated high-sensitivity CRP in serum and detectable TNF-α in milk among women with very low milk supply as compared to women with sufficient milk production, indicating chronic inflammation in the women with very low milk supply. In that study, lower milk production in women with higher inflammation was hypothesized to be driven by suppressed mammary LPL activity, reducing LCFAs lipolysis of serum triglyceride in the mammary gland. In line with this hypothesis, the investigators reported a very strong correlation between LCFA profiles in serum and milk in the control group, but no correlation in the very low milk supply group, which may be explained by downregulated LPL activity in the mammary gland of that group (20). Furthermore, in a murine model, lactation failure occurred more frequently in diet-induced obese mice compared with lean mice. Obese lactating mice had lower concentrations of protein in their milk in comparison to lean lactating mice; however, fat concentration of milk was not reported in the study. Also, the authors reported significantly upregulated expression of inflammatory markers Tnf, Csf1, and Il6, and increased apoptosis and involution of the mammary epithelial cell in obese mice as compared to controls (57).

### Limitations and Strengths

The present study has its limitations. The generalizability of our findings may be limited due to homogeneity of the study participants regarding breastfeeding status and sociodemographic characteristics: Enrollment in the GEHM Study required intention to exclusively breastfeed for the three months and providing at least 75% of feedings as breast milk at week 4 postpartum, and the predominance of participants were non-Hispanic White women. Further, BMI is not an ideal indicator of maternal metabolic health (58–60), and we could not assess biological inflammatory markers in serum to confirm chronic inflammation status of mothers with elevated BMI. We were also limited to the measurement of one inflammatory marker in milk at one time point. We selected CRP because of its ubiquitous associations with low-grade inflammation, and we selected milk samples from week 4 postpartum to maximize sample size. In addition, it would have been ideal to also obtain maternal serum fatty acid profiles to confirm differences in transfer of LCFAs from blood to milk based on maternal diet quality and BMI.

Aside from these limitations, this study has certain strengths. The original GEHM study design was of high quality and included weekly phone surveillance, biweekly study visits, standardized milk collection procedures, and maternal 24-hour diet recalls based on three randomly selected days. Given the scarcity of research on the diet of lactating women in the U.S., especially in Midwestern settings, we believe that our study addresses a critical gap in the literature. We included 40 dietary parameters out of 45 available DII parameters, which strongly supports characterization of dietary inflammatory potential. Another strength of the study is our novel examination of the joint association of maternal BMI and diet quality on milk composition and breastfeeding duration. Furthermore, performing validity and optimization experiments in conjunction with a high-sensitivity ELISA kit to quantify CRP in human milk samples enabled us to examine predictors of milk CRP across a much broader range of values that previously reported for mature milk samples, strengthening the accuracy of our results.

### Conclusions

Our study showed that milk CRP concentrations at four weeks postpartum were lower in women with both a higher quality diet and a normal BMI. We also found that higher prepregnancy BMI and a proinflammatory dietary pattern were associated with lower relative concentrations of milk LCFA and lower EBF duration to 6 weeks. Based on our findings, we recommend that future studies include assessment of maternal serum inflammatory markers and lipid profile, as well as multiple variables that contribute to low-grade inflammation during lactation, including diet quality and measures of maternal adiposity. Further, our data suggest that all cytokine assays used in human milk research should adapted and validated for use in the human milk matrix.

## Data Availability

All data produced in the present study are available upon reasonable request to the authors.

## Abbreviations used

BMI: body mass index
CRP: C-reactive protein
DII: dietary inflammatory index
EBF: exclusive breastfeeding
GEHM: Global Exploration of Human Milk
LCFA: long-chain fatty acid
MCFA: mid-chain fatty acid
WHO: World Health Organization
NDSR: Nutrition Data Systems for Research.

## Notes

### Competing Interest Statement

The authors have declared no competing interest.

### Funding Statement

University of Cincinnati University Research Council (URC) Graduate Student Stipend and Research Cost Program for Faculty-Student Collaboration funded HC in Summer 2022 to perform laboratory analysis; no conflicts of interest. KAD is an employee of Danone Global Research & Innovation Center. Reckitt/Mead Johnson Nutrition funded GEHM milk sample collection but study design, sample analysis, interpretation of results, and writing of the manuscript were fully at the discretion of the authors. NAM and SDM are employees of the Reckitt/Mead Johnson Nutrition. HC, ALM, LNR are funded by NICHD 1R01HD109915.

### Author Declarations

Ethics committee/IRB of Cincinnati Children's Hospital Medical Center gave ethical approval for this work

## References

1. Ballard O, Morrow AL. Human milk composition: nutrients and bioactive factors. Pediatr Clin North Am. 2013;60(1):49–74. doi: 10.1016/j.pcl.2012.10.002.

2. Gila-Diaz A, Arribas SM, Algara A, Martin-Cabrejas MA, Lopez de Pablo AL, Saenz de Pipaon M, Ramiro-Cortijo D. A Review of Bioactive Factors in Human Breastmilk: A Focus on Prematurity. Nutrients. 2019;11(6). doi: 10.3390/nu11061307.

3. Dietz WH. Critical periods in childhood for the development of obesity. Am J Clin Nutr. 1994;59(5):955–9. doi: 10.1093/ajcn/59.5.955.

4. Gillman MW. Early infancy as a critical period for development of obesity and related conditions. Nestle Nutr Workshop Ser Pediatr Program. 2010;65:13–20; discussion -4. doi: 10.1159/000281141.

5. Gillman MW. The first months of life: a critical period for development of obesity. Am J Clin Nutr. 2008;87(6):1587–9. doi: 10.1093/ajcn/87.6.1587.

6. Nommsen LA, Lovelady CA, Heinig MJ, Lonnerdal B, Dewey KG. Determinants of energy, protein, lipid, and lactose concentrations in human milk during the first 12 mo of lactation: the DARLING Study. Am J Clin Nutr. 1991;53(2):457–65. doi: 10.1093/ajcn/53.2.457.

7. Kugananthan S, Gridneva Z, Lai CT, Hepworth AR, Mark PJ, Kakulas F, Geddes DT. Associations between Maternal Body Composition and Appetite Hormones and Macronutrients in Human Milk. Nutrients. 2017;9(3). doi: 10.3390/nu9030252.

8. Leghi GE, Netting MJ, Middleton PF, Wlodek ME, Geddes DT, Muhlhausler ABS. The impact of maternal obesity on human milk macronutrient composition: A systematic review and meta-analysis. Nutrients. 2020;12(4). doi: 10.3390/nu12040934.

9. Dingess KA, Valentine CJ, Ollberding NJ, Davidson BS, Woo JG, Summer S, et al. Branched-chain fatty acid composition of human milk and the impact of maternal diet: the Global Exploration of Human Milk (GEHM) Study. Am J Clin Nutr. 2017;105(1):177–84. doi: 10.3945/ajcn.116.132464.

10. Calder PC, Ahluwalia N, Brouns F, Buetler T, Clement K, Cunningham K, et al. Dietary factors and low-grade inflammation in relation to overweight and obesity. Br J Nutr. 2011;106 Suppl 3:S5–78. doi: 10.1017/S0007114511005460.

11. Ellulu MS, Patimah I, Khaza’ai H, Rahmat A, Abed Y. Obesity and inflammation: the linking mechanism and the complications. Arch Med Sci. 2017;13(4):851–63. doi: 10.5114/aoms.2016.58928.

12. Park HS, Park JY, Yu R. Relationship of obesity and visceral adiposity with serum concentrations of CRP, TNF-alpha and IL-6. Diabetes Res Clin Pract. 2005;69(1):29–35. doi: 10.1016/j.diabres.2004.11.007.

13. Esposito K, Marfella R, Ciotola M, Di Palo C, Giugliano F, Giugliano G, D’Armiento M, D’Andrea F, Giugliano D. Effect of a mediterranean-style diet on endothelial dysfunction and markers of vascular inflammation in the metabolic syndrome: a randomized trial. JAMA. 2004;292(12):1440–6. doi: 10.1001/jama.292.12.1440.

14. Esmaillzadeh A, Kimiagar M, Mehrabi Y, Azadbakht L, Hu FB, Willett WC. Dietary patterns and markers of systemic inflammation among Iranian women. J Nutr. 2007;137(4):992–8. doi: 10.1093/jn/137.4.992.

15. Chrysohoou C, Panagiotakos DB, Pitsavos C, Das UN, Stefanadis C. Adherence to the Mediterranean diet attenuates inflammation and coagulation process in healthy adults: The ATTICA Study. J Am Coll Cardiol. 2004;44(1):152–8. doi: 10.1016/j.jacc.2004.03.039.

16. Serrano-Martinez M, Palacios M, Martinez-Losa E, Lezaun R, Maravi C, Prado M, Martinez JA, Martinez-Gonzalez MA. A Mediterranean dietary style influences TNF-alpha and VCAM-1 coronary blood levels in unstable angina patients. Eur J Nutr. 2005;44(6):348–54. doi: 10.1007/s00394-004-0532-9.

17. Dawod B, Marshall JS. Cytokines and Soluble Receptors in Breast Milk as Enhancers of Oral Tolerance Development. Front Immunol. 2019;10:16. doi: 10.3389/fimmu.2019.00016.

18. Sims CR, Lipsmeyer ME, Turner DE, Andres A. Human milk composition differs by maternal BMI in the first 9 months postpartum. Am J Clin Nutr. 2020;112(3):548–57. doi: 10.1093/ajcn/nqaa098.

19. Whitaker KM, Marino RC, Haapala JL, Foster L, Smith KD, Teague AM, et al. Associations of Maternal Weight Status Before, During, and After Pregnancy with Inflammatory Markers in Breast Milk. Obesity (Silver Spring). 2017;25(12):2092–9. doi: 10.1002/oby.22025.

20. Walker RE, Harvatine KJ, Ross AC, Wagner EA, Riddle SW, Gernand AD, Nommsen-Rivers LA. Fatty Acid Transfer from Blood to Milk Is Disrupted in Mothers with Low Milk Production, Obesity, and Inflammation. J Nutr. 2023;152(12):2716–26. doi: 10.1093/jn/nxac220.

21. Li R, Fein SB, Chen J, Grummer-Strawn LM. Why mothers stop breastfeeding: mothers’ self-reported reasons for stopping during the first year. Pediatrics. 2008;122 Suppl 2:S69–76. doi: 10.1542/peds.2008-1315i.

22. Odom EC, Li R, Scanlon KS, Perrine CG, Grummer-Strawn L. Reasons for earlier than desired cessation of breastfeeding. Pediatrics. 2013;131(3):e726–32. doi: 10.1542/peds.2012-1295.

23. Demmelmair H, Koletzko B. Lipids in human milk. Best Pract Res Clin Endocrinol Metab. 2018;32(1):57–68. doi: 10.1016/j.beem.2017.11.002.

24. Neville MC, Picciano MF. Regulation of milk lipid secretion and composition. Annu Rev Nutr. 1997;17:159–83. doi: 10.1146/annurev.nutr.17.1.159.

25. Shivappa N, Steck SE, Hurley TG, Hussey JR, Hebert JR. Designing and developing a literature-derived, population-based dietary inflammatory index. Public Health Nutr. 2014;17(8):1689–96. doi: 10.1017/S1368980013002115.

26. Shivappa N, Hebert JR, Rietzschel ER, De Buyzere ML, Langlois M, Debruyne E, Marcos A, Huybrechts I. Associations between dietary inflammatory index and inflammatory markers in the Asklepios Study. Br J Nutr. 2015;113(4):665–71. doi: 10.1017/S000711451400395X.

27. Ren Z, Zhao A, Wang Y, Meng L, Szeto IM, Li T, Gong H, Tian Z, Zhang Y, Wang P. Association between Dietary Inflammatory Index, C-Reactive Protein and Metabolic Syndrome: A Cross-Sectional Study. Nutrients. 2018;10(7). doi: 10.3390/nu10070831.

28. Vahid F, Shivappa N, Faghfoori Z, Khodabakhshi A, Zayeri F, Hebert JR, Davoodi SH. Validation of a Dietary Inflammatory Index (DII) and Association with Risk of Gastric Cancer: a Case-Control Study. Asian Pac J Cancer Prev. 2018;19(6):1471–7. doi: 10.22034/APJCP.2018.19.6.1471.

29. Wirth MD, Burch J, Shivappa N, Violanti JM, Burchfiel CM, Fekedulegn D, et al. Association of a dietary inflammatory index with inflammatory indices and metabolic syndrome among police officers. J Occup Environ Med. 2014;56(9):986–9. doi: 10.1097/JOM.0000000000000213.

30. Zhang Z, Wu Y, Zhong C, Zhou X, Liu C, Li Q, et al. Association between dietary inflammatory index and gestational diabetes mellitus risk in a prospective birth cohort study. Nutrition. 2021;87–88:111193. doi: 10.1016/j.nut.2021.111193.

31. Yang Y, Kan H, Yu X, Yang Y, Li L, Zhao M. Relationship between dietary inflammatory index, hs-CRP level in the second trimester and neonatal birth weight: a cohort study. J Clin Biochem Nutr. 2020;66(2):163–7. doi: 10.3164/jcbn.19-100.

32. Sen S, Rifas-Shiman SL, Shivappa N, Wirth MD, Hebert JR, Gold DR, Gillman MW, Oken E. Dietary Inflammatory Potential during Pregnancy Is Associated with Lower Fetal Growth and Breastfeeding Failure: Results from Project Viva. J Nutr. 2016;146(4):728–36. doi: 10.3945/jn.115.225581.

33. Kant AK, Graubard BI. Secular trends in regional differences in nutritional biomarkers and self-reported dietary intakes among American adults: National Health and Nutrition Examination Survey (NHANES) 1988-1994 to 2009-2010. Public Health Nutr. 2018;21(5):927–39. doi: 10.1017/S1368980017003743.

34. Cusack LK, Smit E, Kile ML, Harding AK. Regional and temporal trends in blood mercury concentrations and fish consumption in women of child bearing Age in the united states using NHANES data from 1999-2010. Environ Health. 2017;16(1):10. doi: 10.1186/s12940-017-0218-4.

35. U.S. Department of Agriculture and U.S. Department of Health and Human Services. Dietary Guidelines for Americans, 2020-2025. In: Americans tEDGf, ed., 2020.

36. Committee. DGA. Scientific Report of the 2020 Dietary Guidelines Advisory Committee: Advisory Report to the Secretary of Agriculture and the Secretary of Health and Human Services. . In: Service. USDoAAR, ed. Washington, DC, 2020.

37. Woo JG, Guerrero ML, Ruiz-Palacios GM, Peng YM, Herbers PM, Yao W, Ortega H, Davidson BS, McMahon RJ, Morrow AL. Specific infant feeding practices do not consistently explain variation in anthropometry at age 1 year in urban United States, Mexico, and China cohorts. J Nutr. 2013;143(2):166–74. doi: 10.3945/jn.112.163857.

38. Suwaydi MA, Gridneva Z, Perrella SL, Wlodek ME, Lai CT, Geddes DT. Human Milk Metabolic Hormones: Analytical Methods and Current Understanding. Int J Mol Sci. 2021;22(16). doi: 10.3390/ijms22168708.

39. Fujimori M, Franca EL, Fiorin V, Morais TC, Honorio-Franca AC, de Abreu LC. Changes in the biochemical and immunological components of serum and colostrum of overweight and obese mothers. BMC Pregnancy Childbirth. 2015;15:166. doi: 10.1186/s12884-015-0574-4.

40. Bligh EG, Dyer WJ. A rapid method of total lipid extraction and purification. Can J Biochem Physiol. 1959;37(8):911–7. doi: 10.1139/o59-099.

41. World Health Organization. Indicators for assessing infant and young child feeding practices. Part I: definition. 2008. accessed Date Accessed)|.

42. World Health Organization. A healthy lifestyle - WHO recommendations [Internet]. 2010; Available from: https://www.who.int/europe/news-room/fact-sheets/item/a-healthy-lifestyle---who-recommendations.

43. Aumeistere L, Ciprovica I, Zavadska D, Andersons J, Volkovs V, Celmalniece K. Impact of Maternal Diet on Human Milk Composition Among Lactating Women in Latvia. Medicina (Kaunas). 2019;55(5). doi: 10.3390/medicina55050173.

44. Petersohn I, Hellinga AH, van Lee L, Keukens N, Bont L, Hettinga KA, Feskens EJM, Brouwer-Brolsma EM. Maternal diet and human milk composition: an updated systematic review. Front Nutr. 2023;10:1320560. doi: 10.3389/fnut.2023.1320560.

45. Wong VW, Ng YF, Chan SM, Su YX, Kwok KW, Chan HM, et al. Positive relationship between consumption of specific fish type and n-3 PUFA in milk of Hong Kong lactating mothers. Br J Nutr. 2019;121(12):1431–40. doi: 10.1017/S0007114519000801.

46. R Core Team. R: A Language and Environment for Statistical Computing. Vienna, Austria: R Foundation for Statistical Computing, 2023.

47. Venables WN, Ripley BD. Modern Applied Statistics with S, Fourth edition. New York: Springer, 2002.

48. Friedman J, Hastie T, Tibshirani R. Regularization Paths for Generalized Linear Models via Coordinate Descent. J Stat Softw. 2010;33(1):1–22.

49. Hebert JR, Shivappa N, Wirth MD, Hussey JR, Hurley TG. Perspective: The Dietary Inflammatory Index (DII)-Lessons Learned, Improvements Made, and Future Directions. Adv Nutr. 2019;10(2):185–95. doi: 10.1093/advances/nmy071.

50. Ramiro-Cortijo D, Herranz Carrillo G, Singh P, Rebollo-Hernanz M, Rodriguez-Rodriguez P, Ruvira S, Martin-Trueba M, Martin CR, Arribas SM. Maternal and Neonatal Factors Modulating Breast Milk Cytokines in the First Month of Lactation. Antioxidants (Basel). 2023;12(5). doi: 10.3390/antiox12050996.

51. Minihane AM, Vinoy S, Russell WR, Baka A, Roche HM, Tuohy KM, et al. Low-grade inflammation, diet composition and health: current research evidence and its translation. Br J Nutr. 2015;114(7):999–1012. doi: 10.1017/S0007114515002093.

52. Dallas DC, Murray NM, Gan J. Proteolytic Systems in Milk: Perspectives on the Evolutionary Function within the Mammary Gland and the Infant. J Mammary Gland Biol Neoplasia. 2015;20(3-4):133–47. doi: 10.1007/s10911-015-9334-3.

53. Brockway MM, Daniel AI, Reyes SM, Gauglitz JM, Granger M, McDermid JM, et al. Human Milk Bioactive Components and Child Growth and Body Composition in the First 2 Years: A Systematic Review. Adv Nutr. 2024;15(1):100127. doi: 10.1016/j.advnut.2023.09.015.

54. Centers for Disease Control and Prevention. Breastfeeding Report Card–– United States, 2008 Internet: https://www.cdc.gov/breastfeeding/pdf/2008breastfeedingreportcard.pdf acceSSed Date Accessed)|.

55. Keyes M, Andrews C, Midya V, Carrasco P, Guxens M, Jimeno-Romero A, et al. Mediators of the association between maternal body mass index and breastfeeding duration in 3 international cohorts. Am J Clin Nutr. 2023;118(1):255–63. doi: 10.1016/j.ajcnut.2023.04.004.

56. Thulier D, Mercer J. Variables associated with breastfeeding duration. J Obstet Gynecol Neonatal Nurs. 2009;38(3):259–68. doi: 10.1111/j.1552-6909.2009.01021.x.

57. Hennigar SR, Velasquez V, Kelleher SL. Obesity-Induced Inflammation Is Associated with Alterations in Subcellular Zinc Pools and Premature Mammary Gland Involution in Lactating Mice. J Nutr. 2015;145(9):1999–2005. doi: 10.3945/jn.115.214122.

58. Somi MH, Nikniaz Z, Ostadrahimi A, Eftekhar Sadat AT, Faramarzi E. Is normal body mass index a good indicator of metabolic health in Azar cohort population? J Cardiovasc Thorac Res. 2019;11(1):53–60. doi: 10.15171/jcvtr.2019.09.

59. Nuttall FQ. Body Mass Index: Obesity, BMI, and Health: A Critical Review. Nutr Today. 2015;50(3):117–28. doi: 10.1097/NT.0000000000000092.

60. Carbone S, Canada JM, Billingsley HE, Siddiqui MS, Elagizi A, Lavie CJ. Obesity paradox in cardiovascular disease: where do we stand? Vasc Health Risk Manag. 2019;15:89–100. doi: 10.2147/VHRM.S168946.

