## Supplementary Materials for "Maternal Diet Quality and BMI as Predictors of Human Milk Composition and Exclusive Breastfeeding Duration"

**Abstract**

**Background:** Poor diet quality and high body mass index (BMI) contribute to inflammation, which may influence human milk composition and exclusive breastfeeding (EBF) duration.

**Objective:** We evaluated maternal diet and prepregnancy BMI as predictors of human milk C-reactive protein (CRP) and long chain fatty acid concentrations (%LCFA), and EBF duration.

**Methods:** We utilized the Global Exploration of Human Milk Study-Cincinnati subset (n=114), where healthy dyads continued follow-up if ≥75% of feeds were breastmilk at 4 weeks postpartum. We computed a Dietary Inflammatory Index (DII) from diet recalls obtained between 4-13 weeks postpartum, where higher score indicates a more proinflammatory diet. Milk CRP and fatty acid analyses were performed on week 4 milk. We compared milk CRP across 4 combinations of DII×BMI using the Kruskal Wallis test, with BMI categorized as normal versus elevated (<25 versus ≥25 kg/m^2^), and DII split at the median. Linear regression was used to examine DII and BMI as predictors of %LCFA. Logistic regression was used to examine DII tertiles and BMI as predictors of EBF duration.

**Results:** Milk CRP concentrations differed across DII×BMI groups (*p*=0.009): the low DII/normal BMI group had the lowest milk CRP (n=30, median [Q1, Q3], 64.3 [38.2, 121.4] ng/mL) versus all other groups (n=70, 124.1 [71.2, 181] ng/mL, *p*=0.022). Lower milk %LCFA was predicted by higher DII score (β±SE = -0.68 ± 0.21, *p*=0.002, n=103) and higher BMI (β±SE = -0.13 ± 0.01, *p*=0.043, n=114). Having the highest DII tertile and elevated BMI lowered the odds of EBF at week 6 (OR [95% CI]: 0.26 [0.07, 0.85]) compared to the referent group (low or medium DII, normal BMI).

**Conclusions:** Milk CRP concentrations were lowest in women with a more anti-inflammatory diet and normal BMI. Both higher BMI and proinflammatory diet predict lower milk %LCFA and lower EBF prevalence at week 6.

**Supplementary Information**

**Dietary Inflammatory Index (DII) Computation**

DII is based on the inflammatory effect score of each parameter multiplied by a normalized participant intake z-score relative to a global database. Calculation of the DII score is explained by Shivappa et al. in detail (1). A global database, created by combining the datasets for dietary intake from different regions of the world determines the “global mean intake” and “global standard deviation” for each parameter.

Z-score for each parameter for each subject is derived by subtracting the “standard global mean intake” from the subject’s average dietary intake of the parameter, and dividing this value by “standard global standard deviation” provided by DII developers in the global database. Raw z-scores were converted into a percentile score, which was multiplied by two, and then “1” was subtracted to make a symmetrical distribution. In order to get a ‘food parameter-specific DII score’, the centered percentile score was multiplied by its respective ‘overall food parameter-specific inflammatory effect score’ provided by the DII developers. Lastly, the overall DII score for each participant was achieved when all of the ‘food parameter-specific DII scores’ are summed. The resulting DII score ranges from −8.87 (anti-inflammatory) to +7.98 (proinflammatory). There is no strict requirement to use all 45 possible parameters. In our analysis, we included 40 dietary parameters out of 45 potential parameters, which were energy, carbohydrate, protein, total fat, fiber, cholesterol, saturated fatty acids, monounsaturated fatty acids, polyunsaturated fatty acids, n-3 fatty acids, n-6 fatty acids (obtained by subtracting weight of n-3 fatty acid intake from the total polyunsaturated fatty acids intake), trans-fat, thiamin, riboflavin, niacin, vitamin B-6, vitamin B-12, vitamin A, vitamin C, vitamin D, vitamin E, folic acid, β-carotene, iron, magnesium, zinc, selenium, caffeine, flavones, flavonols, flavan-3-ols, flavanones, anthocyanidins, onion, garlic, ginger, pepper, green/black tea, thyme/oregano, and rosemary. Of these 40 parameters, 34 were available via NDSR 2013, and 7 additional parameters (onion, garlic, ginger, pepper, green/black tea, thyme/oregano, and rosemary) were obtained from scanning the raw dietary data files. We did not include eugenol, turmeric, saffron, isoflavones, alcohol due to lack of data for these parameters. For the majority of nutrients and phytochemicals, the same unit of measurement was used in the NDSR and DII scoring system. However, for vitamin A and folic acid we needed to convert the units used in the NDSR to align with the DII. Briefly, Retinol Equivalents (RE) are used in the DII algorithm, but the NDSR analysis reports separately for total retinol and betacarotene. Therefore, we estimated RE using the formula provided in the NDSR User Manual 2021, in which RE = total retinol (mcg) x (beta-carotene equivalents (mcg)/6). For folic acid, dietary folate equivalents (DFE) as reported in the NDSR were multiplied by 0.6 to estimate folic acid equivalents to align with the DII algorithm (2). In the DII algorithm, the weights of some foods are based on dry weight equivalents, such as spices, tea, and herbs, while the weights of all vegetables and ginger are based on the fresh version. For seven foods in the dietary recall records, the version (dried versus fresh) did not align with the version used in the DII. Therefore, in these cases we converted intake amount recorded in the dietary recall to align with the DII scoring system. Specifically, we used the following factors to convert dried intake to fresh weight equivalents: onion powder or dehydrated onion flakes x 9.0 (3), garlic powder x 4.0 (3), dried chili pepper x 7.74 (4, 5), and ground ginger x 4.29 (6-8). In the DII algorithm, black and green tea are based on dry weight. Therefore, we estimated the dry weight equivalent of tea using the equivalent of 1 gram dried leaf tea per 100 ml of tea beverage consumed (9). In the case of sweetened teas, we subtracted the weight of added sugar from the total beverage intake before calculating the dry weight equivalent. In the DII algorithm, thyme and oregano are combined and based on dry weights. All study participants reported dried versions of these herbs; thus, we summed the intake of both herbs for each participant.

**Supplemental Table 1.** Coefficient of variation for human milk CRP quantification with high-sensitivity ELISA kits at optimal dilution

| **Replicate Number** | **Dilution Factor** | **Optical Density** | **Concentration of CRP (ng/mL)** | **Concentration of CRP x Dilution Factor (ng/mL)** |
| --- | --- | --- | --- | --- |
| **1** | 1:4 | 0.548 | 27.22 | 136.1 |
| **2** | 1:4 | 0.566 | 28.14 | 140.7 |
| **3** | 1:4 | 0.566 | 28.14 | 140.7 |
| **4** | 1:4 | 0.561 | 27.89 | 139.5 |
| **Mean** | 1:4 | 0.560 | 27.85 | 139.3 |
| **SD** | 1:4 | 0.009 | 0.44 | 2.2 |
| **CV %** | 1:4 | 1.520 | 1.56 | 1.6 |

Intra-assay precision was assessed by computing the coefficient of variation (CV %) of the replicates (n = 4) at the appropriate dilution (1:4). Corrected concentrations of CRP in milk was reported after multiplying with dilution factor. ng/mL, nanogram per mililiter; SD, standard deviation; CV %, coefficient of variation.

**Supplemental Table 2.** Spiked recovery values for CRP quantification in human milk with ELISA at 1:4 dilution

| **Number of Replicate** | **Optical Density** | **Amount of CRP standard added (ng)** | **CRP concentration of well (ng/mL)** | **Recovery %** |
| --- | --- | --- | --- | --- |
| **1** | 0.89 | 0.625 | 25.2 | 101.9 |
| **2** | 0.88 | 0.625 | 24.9 | 99.4 |
| **3** | 0.87 | 0.625 | 24.8 | 98.4 |
| **4** | 0.82 | 0.625 | 23.7 | 89.9 |
| **5** | 0.87 | 0.625 | 24.7 | 97.8 |
| **6** | 0.87 | 0.625 | 24.9 | 99.1 |
| **7** | 0.84 | 0.625 | 24.1 | 92.7 |
| **8** | 0.84 | 0.625 | 24.0 | 92.2 |
| **9** | 0.83 | 0.625 | 23.9 | 91.0 |
| **10** | 0.89 | 0.625 | 25.2 | 101.7 |
| **11** | 0.99 | 0.625 | 27.5 | 120.2 |
| **12** | 0.94 | 0.625 | 26.4 | 111.1 |
| **13** | 0.93 | 0.625 | 26.1 | 108.6 |
| **14** | 0.89 | 0.625 | 25.3 | 102.4 |
| **Mean** | 0.88 | 0.625 | 25.1 | 100.5 |
| **SD** | 0.048 | 0 | 1.1 | 8.4 |
| **CV %** | 5.41 | 0 | 4.2 | 8.4 |

Spike recovery was determined by spiking 1:4 diluted skimmed milk samples with CRP (n = 14) standard at the concentration of 25 ng/mL and percent recovery calculated as mean ± SD with CV%.

**Supplemental Table 3.** Summary of Maternal and Infant Characteristics

| Maternal and Infant Characteristics^1^ | Summary statistics |
| --- | --- |
| Total DII score | 0.19 [-1.50, 1.27] |
| Prepregnancy BMI, kg/m^2^ | 25.60 [22.77, 31.12] |
| Maternal delivery age, y | 31.74 ± 5.09 |
| Maternal race  Black  White  Others | 13 (12.04)  93 (86.11)  2 (1.85) |
| Maternal education  High school graduate  College degree  Post-college education | 5 (4.63)  84 (77.78)  19 (17.59) |
| EBF duration, d | 99.5 [51.75, 139.75] |
| BF duration, d | 366.5 [199.8, 487.2] |
| Infant sex, male | 50 (46.30) |
| Infant gestational age, wk | 39.46 ± 1.04 |
| Infant birthweight, kg | 3.57 ± 0.43 |

^1^ Values are mean ± SD or median [Quartile 1, Quartile 3] for parametric and non-parametric continuous variables, respectively or n (%) for frequency (proportion) of categorical variables, n = 108. DII, dietary inflammatory index; BMI, body mass index; kg/m^2^, kilograms per meter square; y, years; d, days; wk, weeks

**Supplemental Figure 1.** Survival model for duration of EBF by week 13 by DII tertiles (A) and groups (B)

A. Kaplan Meier survival curve for EBF proportion by week 13 postpartum across DII tertiles

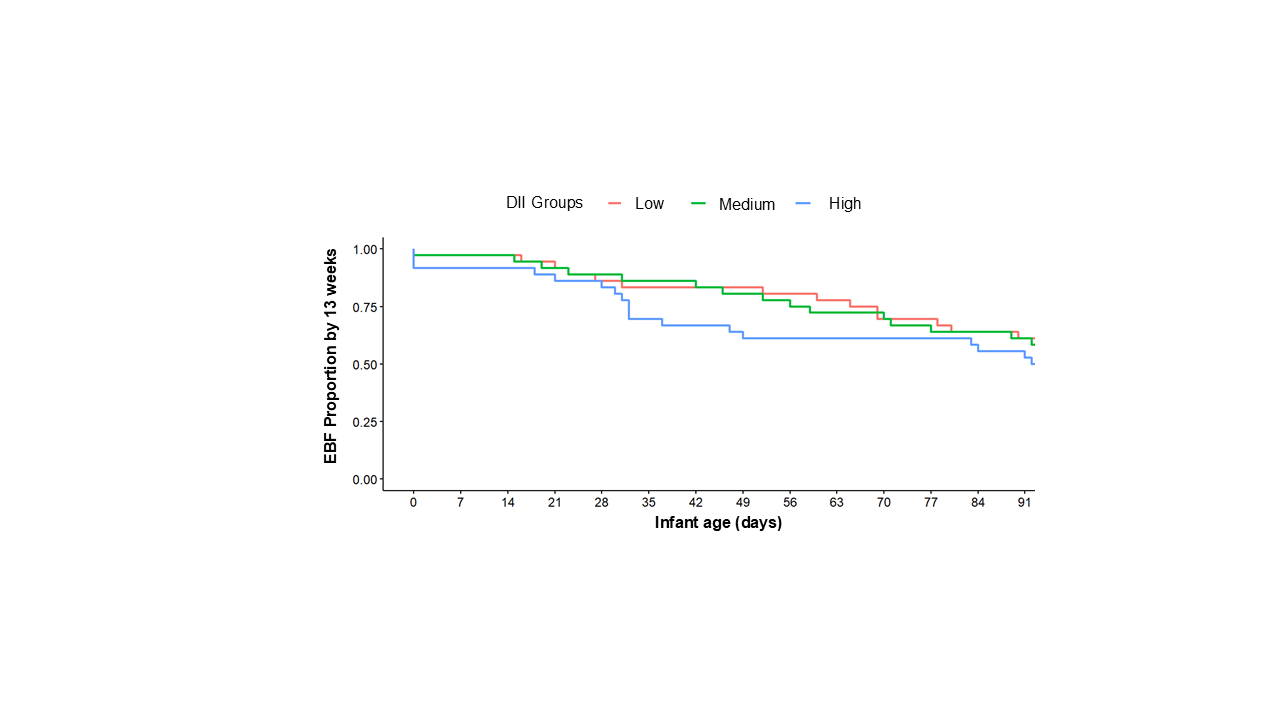

B. Kaplan Meier survival curve for EBF proportion by week 13 postpartum across DII groups

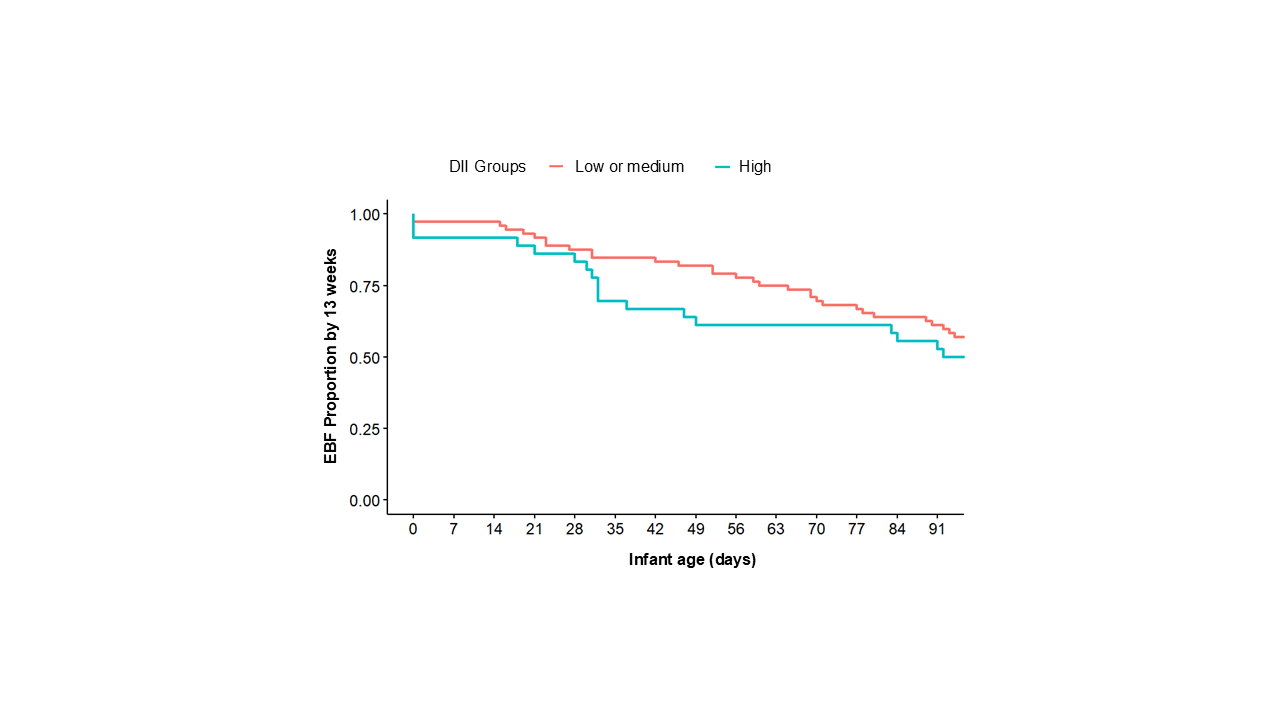

Proportion of EBF by 13 weeks postpartum was compared across DII tertiles (A) and binary grouped DII groups (B) in non-censored observations. EBF, Exclusive breastfeeding; DII, dietary inflammatory index

**Supplemental Table 4.** Associations of maternal DII score and prepregnancy BMI with milk CRP levels (n = 100)

| Maternal Characteristics | Human Milk CRP Levels (ng/mL) | |
| --- | --- | --- |
|  | β ± SE | *p* value |
| BMI | 5.25 ± 1.42 | 0.0004 |
| DII Score | -5.67 ± 4.36 | 0.20 |

Multivariate linear regression analysis to evaluate the relationship between BMI and DII to predict milk CRP levels. CRP, C-reactive protein; BMI, body mass index; DII, dietary inflammation index; β ± SE, beta coefficient ± standard error

**Supplementary Table 5.** Sensitivity analysis for the DII and relative LCFAs concentration of milk ^1^

| Milk fatty acid groups^2^ | Relative concentrations^3^ (Mean ± SD) | Correlation with DII  (n= 103) | |
| --- | --- | --- | --- |
|  |  | B coefficient + Standard error | P value |
| MCFA | 12.1 (3.7) | 0.44 ± 0.18 | 0.014 |
| C16 | 22.5 (2.9) | 0.12 ± 0.15 | 0.416 |
| LCFA | 65.4 (4.6) | -0.57 ± 0.23 | 0.016 |

^1^ To account for the effects of dietary fatty acids employed in the computation of DII score on milk fatty acid profile, we removed 3 DII parameters consisting of dietary LCFAs (omega 3 fatty acids, omega 6 fatty acids, and poly-unsaturated fatty acids). Range of DII score without omega 3, omega 6, and polyunsaturated fatty acids parameters was from -3.45 to +6.64, with median [Q1, Q3] value of 0.27 [-0.87, 1.52].

^2^ Fatty acid groups were created based on fatty acid carbon chain length: medium chain fatty acids (MCFA) are < 16 carbons, C16 = 16 carbon fatty acids, and long chain fatty acids (LCFA) are > 16 carbons.

Then we evaluated the association of DII with relative concentrations of milk fatty acid groups using the total DII score computed with 37 parameters. Despite removing the DII parameter with LCFAs, we observed the negative association between DII and %LCFA in milk.

**References**

1.	Shivappa N, Steck SE, Hurley TG, Hussey JR, Hebert JR. Designing and developing a literature-derived, population-based dietary inflammatory index. Public Health Nutr. 2014;17(8):1689-96. doi: 10.1017/S1368980013002115.

2.	Nutrition Data Systems for Research. *Nutrition Data Systems for Research User Manual* [Internet]. 2021; Available from: http://www.ncc.umn.edu/products/ndsr-user-manual/.

3.	U.S. Department of Agriculture. *Economic Research Service in cooperation with the Agricultural Marketing Service, the Agricultural Research Service, and the National Agricultural Statistics Service. Weights, Measures, and Conversion Factors for Agricultural Commodities and Their Products* [Internet]. Agricultural Handbook No. 697, 1992; Available from: https://www.ers.usda.gov/webdocs/publications/41880/33132_ah697_002.pdf.

4.	U.S. Department of Agriculture. *Food Data Central: Peppers, hot chili, red, raw* [Internet]. Available from: https://fdc.nal.usda.gov/fdc-app.html#/food-details/170106/nutrients.

5.	U.S. Department of Agriculture. *Food Data Central: Peppers, hot chili, sundried* [Internet]. Available from: https://fdc.nal.usda.gov/fdc-app.html#/food-details/168570/nutrients.

6.	U.S. Department of Agriculture. *Food Data Central: Ginger root, raw* [Internet]. Available from: https://fdc.nal.usda.gov/fdc-app.html#/food-details/169231/nutrients.

7.	U.S. Department of Agriculture. *Food Data Central: Spices, ginger, ground* [Internet]. Available from: https://fdc.nal.usda.gov/fdc-app.html#/food-details/170926/nutrients.

8.	Bowman SA MC, Carlson JL, Clemens JC, Lin B-H, and Moshfegh AJ. Food Intakes Converted to Retail Commodities Databases: 2003-08: Methodology and User Guide. US Department of Agriculture, Agricultural Research Service, Beltsville, MD, and US Department of Agriculture, Economic Research Service, Washington, DC. 2013.

9.	Khan N, Mukhtar H. Tea polyphenols for health promotion. Life Sci. 2007;81(7):519-33. doi: 10.1016/j.lfs.2007.06.011.
